# LABELLING HUMAN KINEMATICS DATA USING CLASSIFICATION MODELS

**DOI:** 10.1101/2022.02.18.22271206

**Authors:** Yuan Shi, Nihir Chadderwala, Ujjwal Ratan

## Abstract

The goal of this study is to develop a classification model that can accurately and efficiently label human kinematics data. Kinematics data provides information about the movement of individuals by placing sensors on the human body and tracking their velocity, acceleration and position in three dimensions. These data points are available in C3D format that contains numerical data transformed from 3D data captured from the sensors. The data points can be used to analyse movements of injured patients or patients with physical disorders. To get an accurate view of the movements, the datasets generated by the sensors need to be properly labelled. Due to inconsistencies in the data capture process, there are instances where the markers have missing data or missing labels. The missing labels are a hindrance in motion analysis as it introduces noise and produces incomplete datapoints of sensor’s positioning in 3 dimensional space. Labelling the data manually introduces substantial effort in the analysis process. In this paper, we will describe approaches to pre-process the kinematics data from its raw format and label the data points with missing markers using classification models.

## Introduction

Biomechanics refer to the study of mechanical laws relating to the movement of living organisms.(1) When applied to humans and combined with kinematics, it helps us capture data which is used to infer a variety of human actions. For example, kinematics data is used to understand human’s movements in each gesture, through providing quantitative data to evaluate each individual’s flexibility(2). It is also used to analyse patients’ movements to track their recovery and assist towards rehabilitation(3). Kinematics data is captured in dedicated labs setup as motion capture studios. In some cases, the sensor data may not be labelled completely. This can happen for a variety of reasons like the sensor not being attached securely or some sensors getting blocked from a camera from certain angles. This introduces noise and missing values in the dataset and poses a challenge for further analysis of this data. Capturing movements of these patients and labelling them can be very tedious and an expensive process. To solve this problem, we describe a method where we use the labelled data from these sensors for training a classification model that can then label the unlabelled sensor records with high accuracy, thereby reducing manual efforts.

The primary objective of this study is to use a classification model to label the sensor data corresponding to the mounted location on the human subject’s body. We took the records with pre-labelled information and built classification models that would identify the right class (sensor label) for the unlabeled dataset. In this study, we firstly clean and standardize the raw C3D files through data transformation functions. We then use the processed data to train 4 different machine learning models to classify the sensor data points into one of the multi-class labels. Lastly, we evaluate the performance of the models to auto generate labels with test dataset. Interpretation and future work is provided at the end of the paper.

## Method

In this section, we go into the details of our method starting with describing the raw dataset, data processing and feature engineering, modelling approaches including the choice of algorithms and the evaluation methods.

### Data Description

For the purposes of this study, we downloaded the raw data from CMU Graphics Lab Motion Capture Database in *C3D* format(4), which is a file format that has been widely used in Biomechanics, Animation and Gait Analysis laboratories to record synchronized *3d* kinematics data. It contains information needed to read, display, and analyse *3d* motion data and additional analog data from force plates, electromyography, accelerometers, and other sensors. The dataset contains numerical data extracted from sensors attached to the human body. It is composed of a list of time-series points. Each point is composed of *x, y* and *z* co-ordinates, time of capturing, frame number and the labeled location. The choice of number of sensors and locations mounted vary in each setting as it subjects to the purpose of specific biomedical study. For more details, readers can refer to CMU Graphics Lab Motion Capture Database (4) to visualize how the sensors are mounted on the human body to capture motion data.

The *C3D* dataset being captured can be converted into a standard dataframe (Table 1) using the *c3d* python library(5).

**Table 1:** C3D Sample File in DataFrame

| time | x | y | z | cam | err | frame | point_label |
| --- | --- | --- | --- | --- | --- | --- | --- |
| 1.0125 | 0 | 0 | 0 | FALSE | FALSE | 1 | R_ASIS |
| 1.0125 | 0 | 0 | 0 | FALSE | FALSE | 1 | L_ASIS |
| 1.0125 | 0 | 0 | 0 | FALSE | FALSE | 1 | SACRUM |
| 1.0125 | 0 | 0 | 0 | FALSE | FALSE | 1 | R_THIGH_1 |
| 1.0125 | 0 | 0 | 0 | FALSE | FALSE | 1 | R_THIGH_2 |

Note: This table captures the structure of a raw C3D file when its converted into a dataframe.

As shown in Table 1, a typical *C3D* dataset has 8 features, correspond to *time*,, *x, y, z, cam, err, frame and point_label*. Here, *time* refers to the capture time of the point starting from 0, *x, y* and *z* correspond to captured location at *x, y* and *z* axis respectively. *cam* suggests if there is any camera observing the sensor. To ensure accuracy of captured data, it is required to ensure that at least one camera is observing a sensor at the indicated time. Otherwise, the data point is advised to be removed from the dataset. *err* suggests whether there is error in capturing the *3d* location at this frame, and *frame* refers to the time frame of the current position with continuous integer starting from 1. *Point_labels* are the labelled targets which indicate the location where the sensor is attached to. In our experiment, we downloaded a collection of C3D files of *subject #26* provided by Qualisys illustrating human gait(6). Among all the collections, we selected all the files titled as hybrid walking motions, containing 5 C3D files in total.

### Data Processing and Feature Engineering

Data processing is an important stage to prepare data for machine learning models. In addition, data processing gives us a chance to gain insight into the data and perform feature engineering. Using C3D python library (5), we were able to extract *x, y* and *z* co-ordinates for each frame of the motion. We followed the following data processing steps :

#### 1) Identify all missing values and convert them to NaN

An initial investigation of the dataset showed that all the missing values captured by sensors have been recorded as 0. As column z has only positive data, while both x and y can have negative, positive and 0 values, we used *z* as reference to identify missing values and convert all the corresponding values from all axis to *NaN* if *z=0*.

#### 2) Removal of missing values

Removal of missing values includes:

a. removal of *sensors* which failed to capture locations with 50% or more of the total frames. After applying the filtering criteria, we were left with 19 labels among the initial 25 labels in total.
b. removal of *frames* with consecutive missingness >3 as too many consecutive missingness will result in bad interpretation during data processing.

#### 3) Interpolation

Polynomial interpolation was later carried out with sensors that contain missing values. We chose polynomial interpolation with 3-degree as it is simple and is still able to approximate complicated curves. As showed in the equation below, we refer to *P(x)* as the polynomial function with degree of 3.

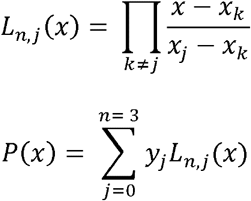

#### 3) Feature Engineering

Feature engineering was conducted on the sensor data based on kinetic understanding. The sensors are attached to human body, each part of human body will move according to a specific trajectory, thus the spatial change of each sensor can be a good indicator of where the sensor is mounted on the human body. Based on this assumption, we included the following features in our training set:

1. Absolute location: *x, y* and *z* values directly generated from the device. Here, *t* refers to the time point of measurement and *x*_*t*_, *y*_*t*_ and *z*_*t*_ refer to the absolute position of respective axis at time *t*.
2. Relative location: the relative rank of each sensor at time point *t*. It is calculated through ranking each point *x, y* and *z* values among all the *x, y* and *z* at each time *t* accordingly. Let’s take *x* for example.

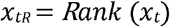
3. Relative change (1-dimensional): the change to the current frame from previous *i* frame(s).

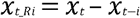
4. Relative change (3-dimenisonal): the change from previous frame to the current frame.

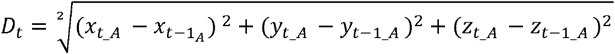

In the classification models, the raw data as absolute location (feature 1) plus time is used in our MLP base and LSTM models, while raw data and enhanced features 1-4 are used in MLP and XGBoost classification.

After feature engineering, the first 4 files are connected together as training dataset and the file 5 is left as testing dataset.

### Modelling Algorithms

Our goal of the experiment was to accurately classify the sensor data into one of the 19 sensor labels in our dataset. We assumed the point_label column as the target variable for this classification task. We selected multilayer perceptron (MLP) base model using scikit-learn, XGBoost and Long Short-Term Memory (LSTM) networks for this work.

### Baseline Model with Multi-Layer Perceptron

We decided to use Multilayer Perceptron (MLP) as our base model (Figure 1) as they are good for both classification and regression problem and can work very well with tabular dataset.

**Figure 1:**
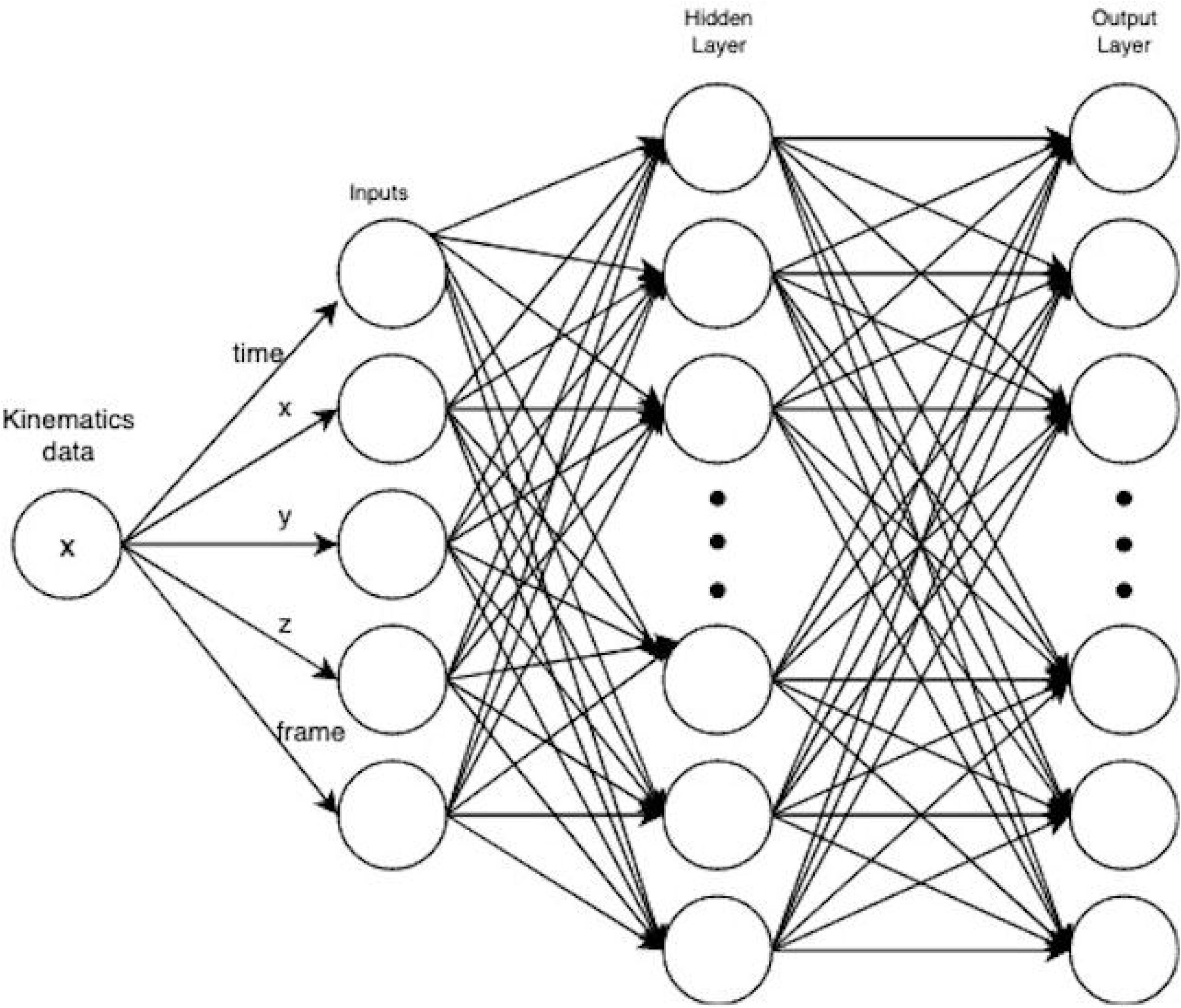
Model topology of MLP baseline. In the basic model, we used raw kinematic data including time, frame, x, y and z as model input, with one hidden layer and output of 23 values corresponding to the 23 unique sensor labels.

MLPs are universal function approximators as shown by Cybenko’s theorem(7), and it is a classical neural network where we have input layer, with one or more hidden layer and an output layer.

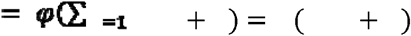

Here, *w* denotes the vector of weights, *x* is the vector of inputs, *b* is the bias and *φ* refers to the non-linear activation function.

Training MLP involves multiple passes on the dataset while at the same time adjusting weights and biases in relation to the error with the goal of reducing the error. MLP model adjusts the weights and biases using a technique called backpropagation. During forward pass, our input vector passes through the input layer and activation function. The result is then compared with the ground truth value to calculate the error using a loss function. In the backward pass, we compute the gradients using stochastic gradient descent algorithm and adjust the weights and biases. Weights and biases are adjusted in order to reduce the error when making classification.

### Enhanced MLP

On top of all the absolute features that we included in MLP baseline model, we further included enhanced features as mentioned in 2.2 data processing section.

As evident from the diagram (Figure 2), additional features are fed into the model as inputs. Features such as frame and time were removed for this model while new features such as changes of position in *x, y* and *z* from previous time point to current time point were added. Training the model using feature engineered data helped improve the model.

**Figure 2:**
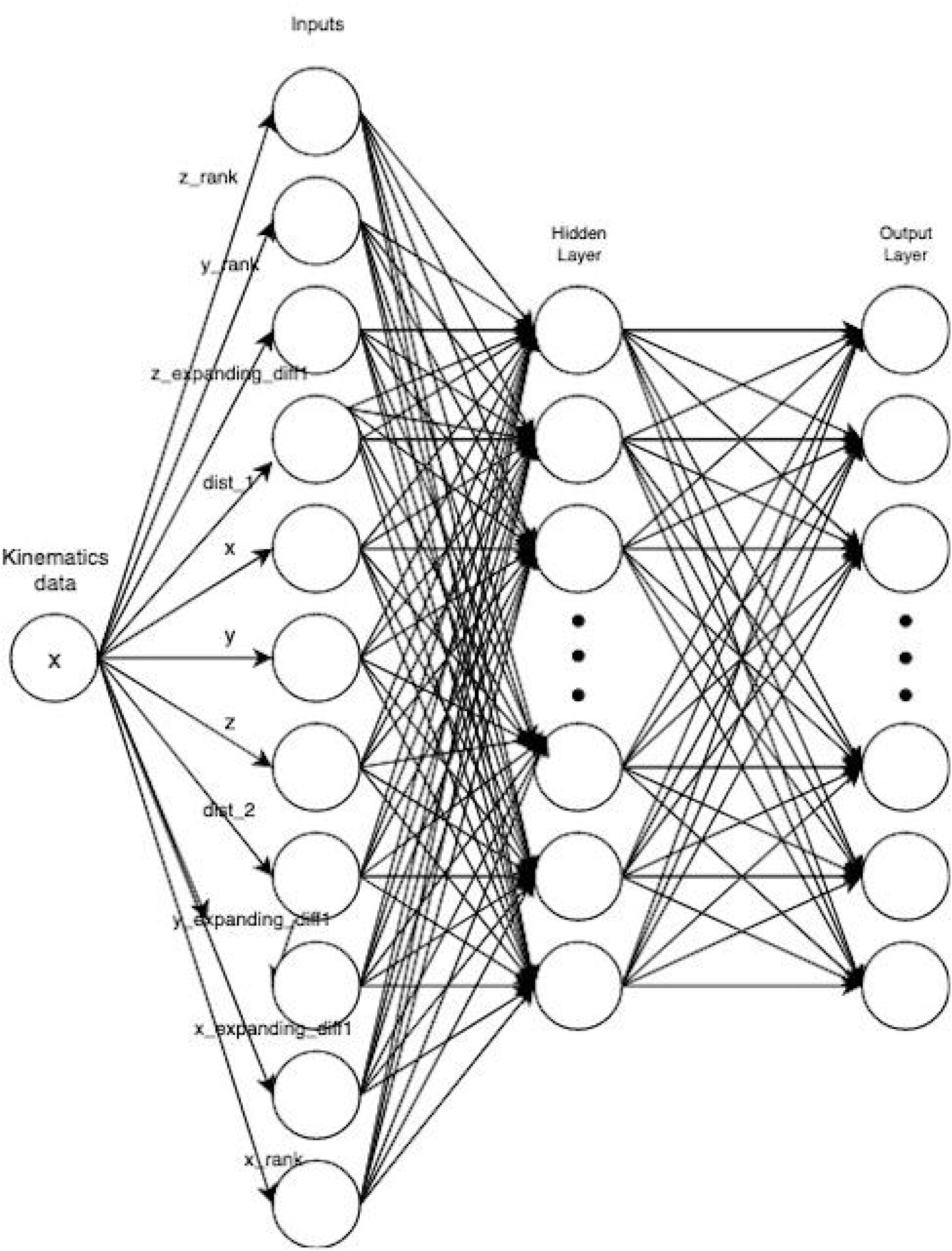
Model topology of MLP enhanced. In this enhanced model, we included features as introduced in method section while retaining model structure as baseline model.

### XGBoost

XGBoost was initially proposed by *Chen and Guestrin* in 2016(8) and it implements machine learning algorithms under the Gradient Boosting framework(9). Specifically, XGBoost is a decision-tree-based ensemble machine learning algorithm based on optimized distributed gradient boosting library and is thus highly efficient, flexible and portable. We followed the default hyperparameters, except for *objective* being changed to *multi:softmax* and the *number_class* being changed to the corresponding 19 classes in the raw C3D dataset.

### LSTM

Long short-term memory (LSTM) is an artificial recurrent neural network (RNN) architecture(10) with a LSTM unit composed of a cell, an input gate, an output gate and a forget gate. For the model topology (Table-2), a sequential model which is a linear stack of layers is used. the first layer is an LSTM layer with memory units and it returns sequence. A dropout layer is applied to avoid overfitting of the model; after that, a dense layer with activation algorithm of *relu* is added, followed by a dense layer with *softmax* function for classification.

**Table 2:**
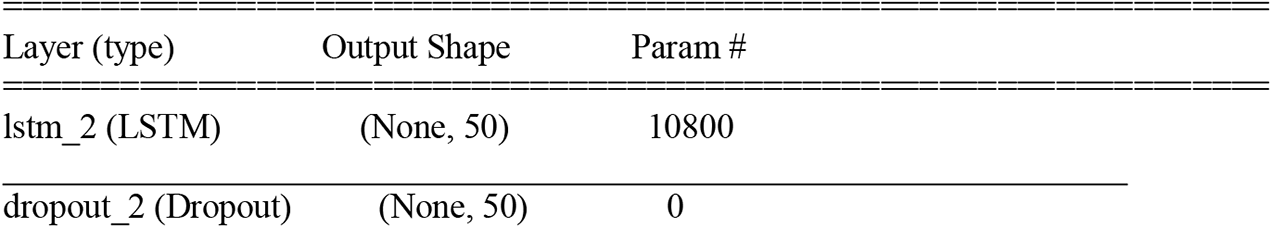
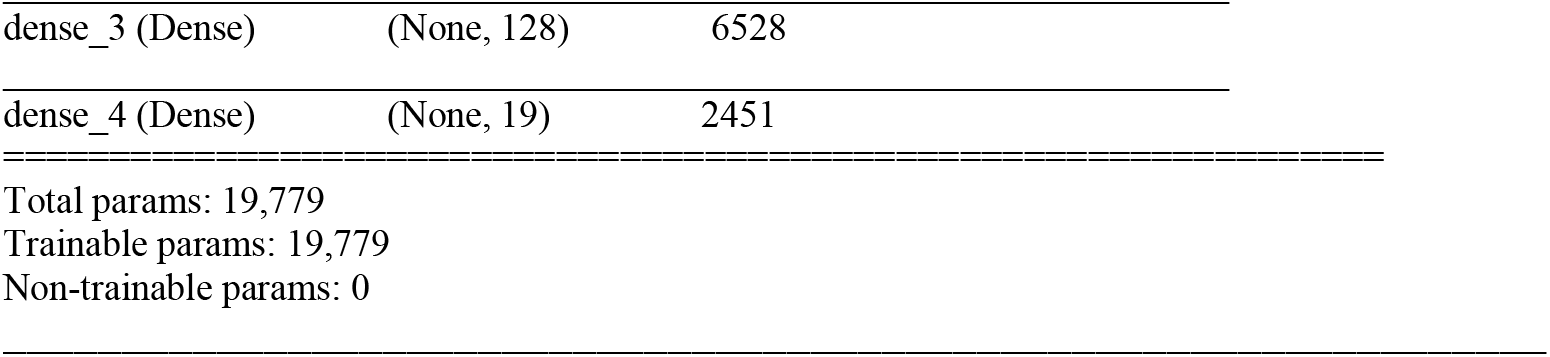
Model topology of LSTM

For the hyperparameters, we set target to maximize *categorical_crossentropy*, with 80 epochs and batch size of 100.

### Model Evaluation Approach

#### F1 score

We are dealing with multi-class classification for a set of time-series data points and the purpose is to classify each set of data into one of the classes. The total dataset is randomly split into training and testing dataset with ratio of 0.8, 0.2. Evaluation is carried out on the testing dataset. As the data labels are not uniformly distributed, we chose the F1 score as the harmonic mean of the precision and recall.

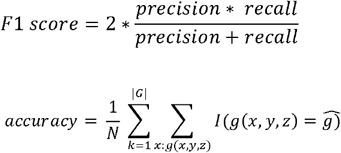

Where *precision* (also called positive predictive value) is the fraction of true positive samples among the predicted true samples, while *recall* (also known as sensitivity) is the fraction of true positive samples among all the positive samples.

#### Confusion matrix

A confusion matrix is also known as an error matrix. It is a table layout that allows visualization of the performance of a supervised learning model. Each row of the matrix represents the instances in an actual class while each column represents the instances in a predicted class, or vice versa.

## Results

Both training dataset and testing dataset were applied with the same data cleaning strategies with polynomial interpolation and all the feature engineering as described in the methodology. After that, we built four models with MLP baseline, MLP enhanced model, XGBoost enhanced model and LSTM using training dataset. The performance was measured on the testing dataset.

### Model Accuracy

The Table 3 below showed performance (F1 score) of the four models with the same testing dataset. As shown in the table, XGBoost showed highest performance with f1 score at 0.94, followed by MLP enhanced (F1 score=0.86), LSTM(F1 score=0.65) and MLP baseline (F1 score=0.64).

**Table 3:** Performance comparison among 4 models

| Model | Algorithm | Feature | F1 score |
| --- | --- | --- | --- |
| MLP baseline | MLP | absolute location, frame | 0.64 |
| MLP enhanced | MLP | absolute location, relative location, changes | 0.86 |
| XGboost enhanced | XGBoost | absolute location, relative location, changes | 0.94 |
| LSTM | LSTM | absolute location | 0.66 |

*Note: This table captures the results of the various algorithms and models used in the evaluation with* *their corresponding F1 scores.*

### Confusion matrix to show agreement between truth and prediction from the model

As XGboost showed highest performance among all the 4 models, we chose predictions from XGboost and use a confusion matrix to demonstrate the agreement between true labels and model output (Figure 3). As illustrated by the side bar, lighter colors refer to higher number while darker colors indicate lower number. In the confusion matrix, we can observe that the matrix diagonal are in lighter color, suggesting high agreement between true labels and predicted labels. Nevertheless, Mislabelling was observed, especially more frequent between *R_HEEL* and *R_MT5*.

**Figure 3:**
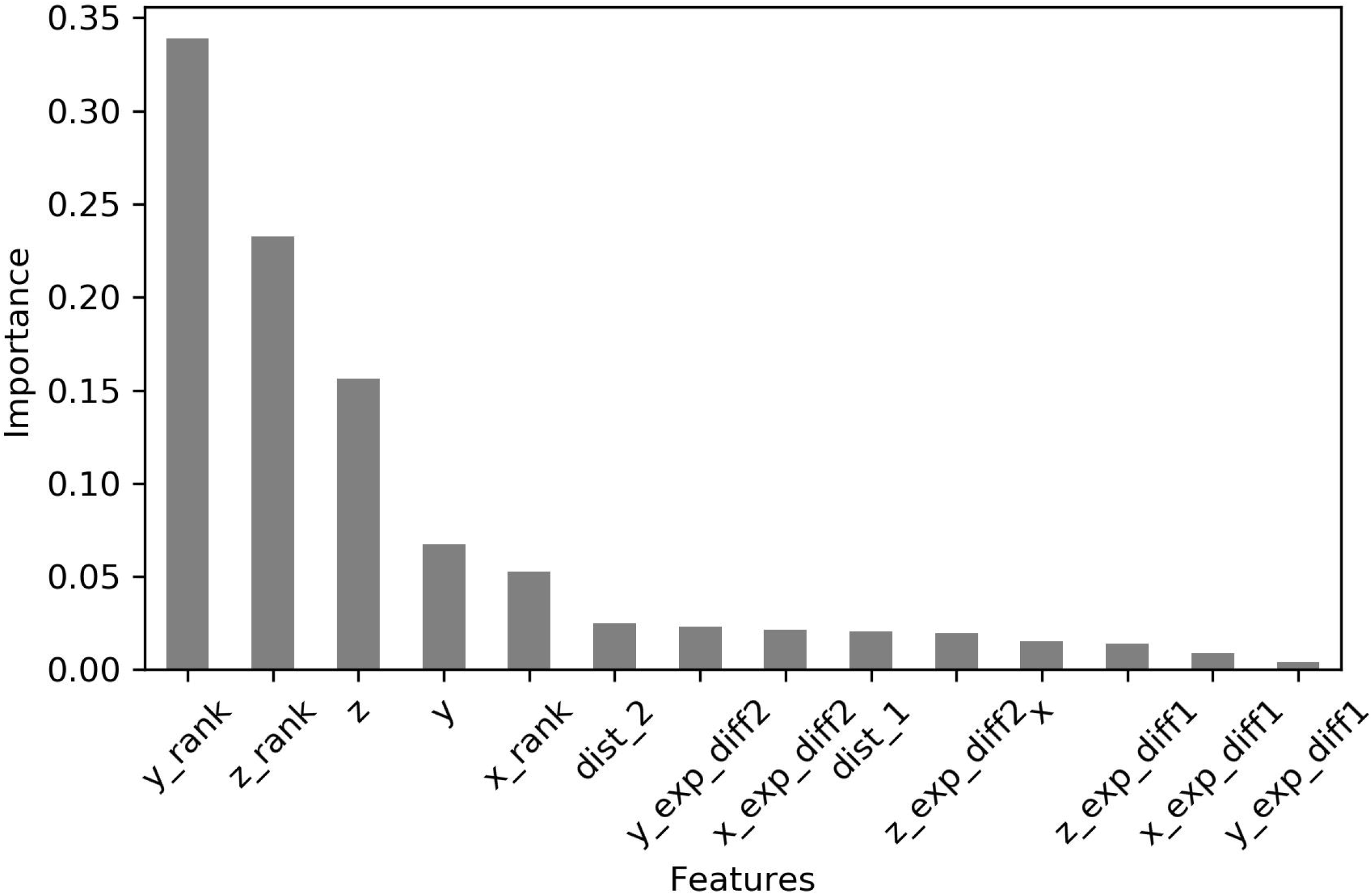
Confusion matrix of true labels and predicted labels from model with XGBoost.

### Feature Importance

As XGboost showed highest accuracy among all the model, we further examined the model by checking feature importance. As we can see in figure 4, it showed that relative location of *y* together with *z* has the highest importance in the modelling, followed by absolute position of z.

**Figure 4:**
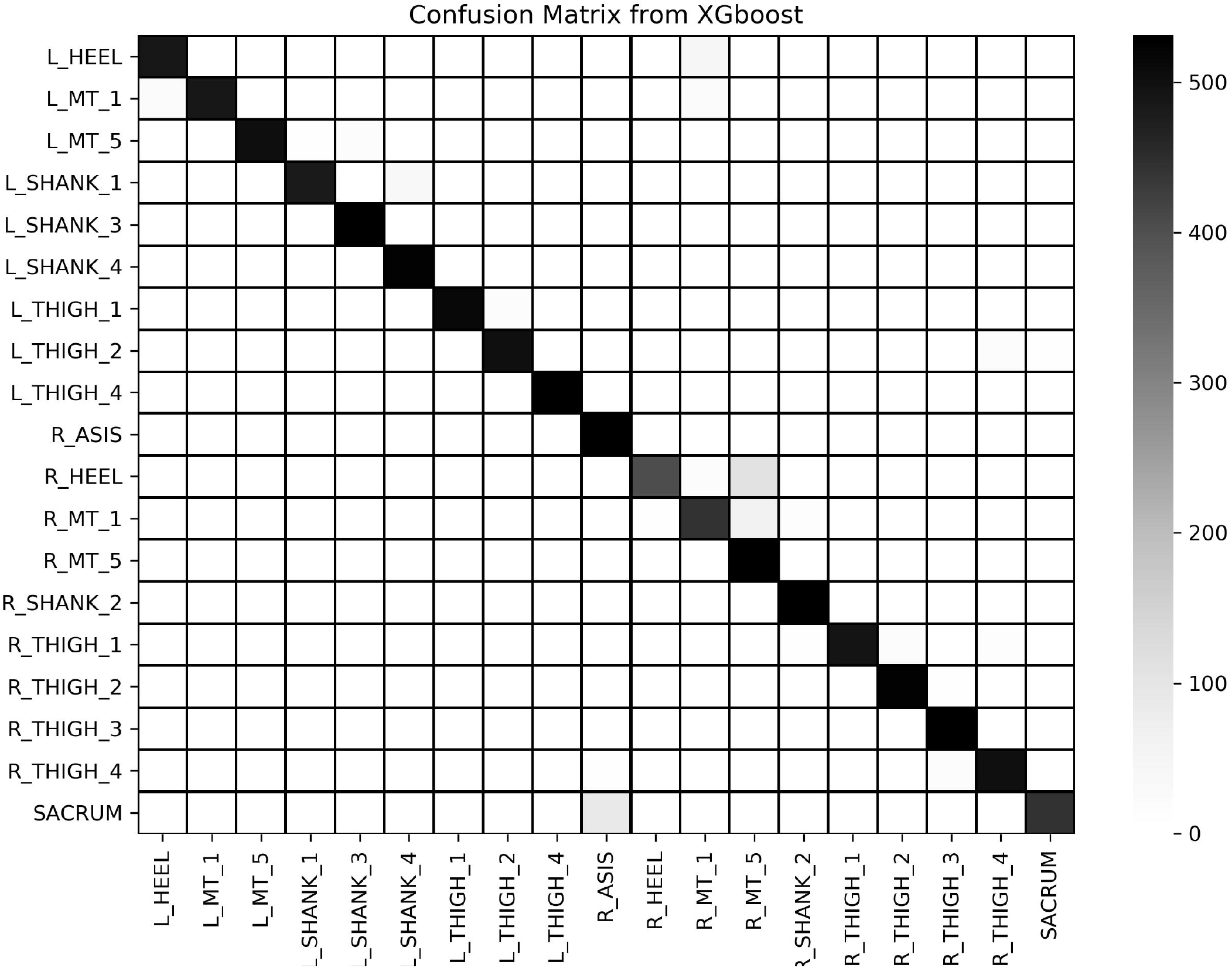
Feature Importance from model with XGBoost.

## Discussion

Capturing human kinematics data requires mounting sensors to pre-defined positions on different parts of the subject’s body, which may lead to missing labels. In current motion capture pipelines, manual validation and labelling is a time consuming and labour intensive postprocessing step and may become a bottleneck for downstream analysis. Prior to this, Holzreiter(11) used neural network to estimate the positions of sorted markers from a shuffled set through pairing up the marker locations with the shuffled set using the nearest neighbour search. Meyer *et al*.(12) estimated the skeletal configuration by least-squares optimization and exploited the skeletal model to automatically label the markers. Besides, Han *et al*.(13) proposed auto-labelling approaches specifically for labelling hands’ kinematics data through keypoint regression problem solved with convolutional neural networks. In addition, Saeed *et al*.(14) used a data-driven approach for automatic labelling through permutation estimation where shuffled markers are ranked based on a pre-defined order.

While the previous work has no labelled data and tried to classify the kinematic data to different locations according to the temporal profile, our work targets to label kinematic data with missing labels. Specifically, the problem we are focusing on is the data quality of the captured data with majority of the data being labelled and a few missing data points due to technical errors. After effective feature engineering and modeling with 3 selected model algorithms, we are able to achieve decent classification outome with F1-score over 90%.

The list of features, model performance and feature importance will vary based on the type of motion being captured by the sensors. In this experiment, we used the motion data of a walking subject, on which over 20 sensors were mounted. Since the motion was horizontal, all sensors moved by similar distance along the *x* axis as the subject moved. As a result, the relative position of *x* did not contribute as much to differentiation of sensors, as compared to *y* and *z* axes, because the sensors were mounted at different hights on the subject’s body. To apply the models to other movements, we need to retrain the model with least effort since the list of features, model performance and feature importance will vary based on the type of motions being captured by the sensors. We also noticed that appropriate feature engineering was essential to get higher accuracy. By using relative locations of the sensors along the *x, y* and *z* axes, we were able to improve the performance of the model by over 31% compared to the baseline model that used just the absolute locations of the sensors. This shows that relative location was the key factor in determining the appropriate sensors.

## Conclusion

We tested different data transformation and machine learning algorithms to develop a multi-class classification model that can label kinematics data with high accuracy. It was also important to apply techniques on kinematics timeseries data as it helped us to improve the model. Calculating the 1-dimensional and 3-dimensional relative change to the frames helped with creation of new features. These techniques improved the model performance considerably. XGBoost model gave the best performance compared to the neural architecture models. The models can be used to accurately label the three-dimensional motion data which can provide insights into movements of a patient with injury or a patient with disability. Analyzing these movements can further help in either creating a recovery plan or an exoskeleton that can aid in recovery.

## Data Availability

All data produced in the present work are contained in the manuscript

https://www.c3d.org/data/Sample26.zip

